# The effect of human mobility and control measures on the COVID-19 epidemic in China

**DOI:** 10.1101/2020.03.02.20026708

**Authors:** Moritz U.G. Kraemer, Chia-Hung Yang, Bernardo Gutierrez, Chieh-Hsi Wu, Brennan Klein, David M. Pigott, open COVID-19 data working group, Louis du Plessis, Nuno R. Faria, Ruoran Li, William P. Hanage, John S. Brownstein, Maylis Layan, Alessandro Vespignani, Huaiyu Tian, Christopher Dye, Simon Cauchemez, Oliver G. Pybus, Samuel V. Scarpino

## Abstract

The ongoing COVID-19 outbreak has expanded rapidly throughout China. Major behavioral, clinical, and state interventions are underway currently to mitigate the epidemic and prevent the persistence of the virus in human populations in China and worldwide. It remains unclear how these unprecedented interventions, including travel restrictions, have affected COVID-19 spread in China. We use real-time mobility data from Wuhan and detailed case data including travel history to elucidate the role of case importation on transmission in cities across China and ascertain the impact of control measures. Early on, the spatial distribution of COVID-19 cases in China was well explained by human mobility data. Following the implementation of control measures, this correlation dropped and growth rates became negative in most locations, although shifts in the demographics of reported cases are still indicative of local chains of transmission outside Wuhan. This study shows that the drastic control measures implemented in China have substantially mitigated the spread of COVID-19.

## Main text

The outbreak of COVID-19 has spread rapidly from its origin in Wuhan, Hubei Province, China (*1*). A range of interventions have been implemented following the detection in late December 2019 of a cluster of pneumonia cases of unknown etiology, and identification of the causative virus SARS-CoV-2 in early January 2020 (*2*). Interventions include improved rates of diagnostic testing, clinical management, rapid isolation of suspected and confirmed cases and, most notably, restrictions on mobility (hereafter called cordon sanitaire) imposed on Wuhan city on 23^rd^ January. Travel restrictions were subsequently imposed on 14 other cities across Hubei Province and partial movement restrictions have been enacted in many cities across China. Initial analysis suggests that the Wuhan cordon sanitaire resulted in an average delay of COVID-19 spread to other cities of 3 days (*3*), but the true extent of the effect of the mobility restrictions and other types of interventions on transmission has not been examined in detail (*4, 5*). Questions remain over how these interventions affected the spread of SARS-CoV-2 to locations outside of Wuhan. We here use real-time mobility data, crowdsourced line-list data of cases with reported travel history, and timelines of reporting changes to identify early shifts in the epidemiological dynamics of the COVID-19 epidemic in China, from an epidemic driven by frequent importations to local transmission.

### Human mobility predicts the spread and size of epidemics in China

As of 1^st^ March 2020, 79,986 cases of COVID-19 have been confirmed in China (**Fig. 1a**) (*10*). Reports of cases in China were mostly restricted to Hubei until 23^rd^ January 2020 (81% of all cases), after which most provinces reported rapid increases in cases (**Fig. 1a**). We build a line list dataset from reported cases in China with information on travel history and demographic characteristics. We note that the majority of early cases (before 23^rd^ January 2020, **Materials and Methods**) reported outside of Wuhan had known travel history to Wuhan (57%) and were distributed across China (**Fig. 1b**), highlighting the importance of Wuhan as the major source. However, testing was initially focused mainly on travelers to Wuhan, potentially biasing our estimates of travel related infections upwards (**Materials and Methods**). Among cases known to have traveled from Wuhan before 23^rd^ January 2020, the time from symptom onset to confirmation was 6.5 days (SD: 4.2, **Fig. S2**), providing opportunity for onward transmission at the destination. More active surveillance brought this interval down to 4.8 days (SD: 3.03, **Fig. S2**) for those who travelled after 23^rd^ January 2020.

**Figure 1:**
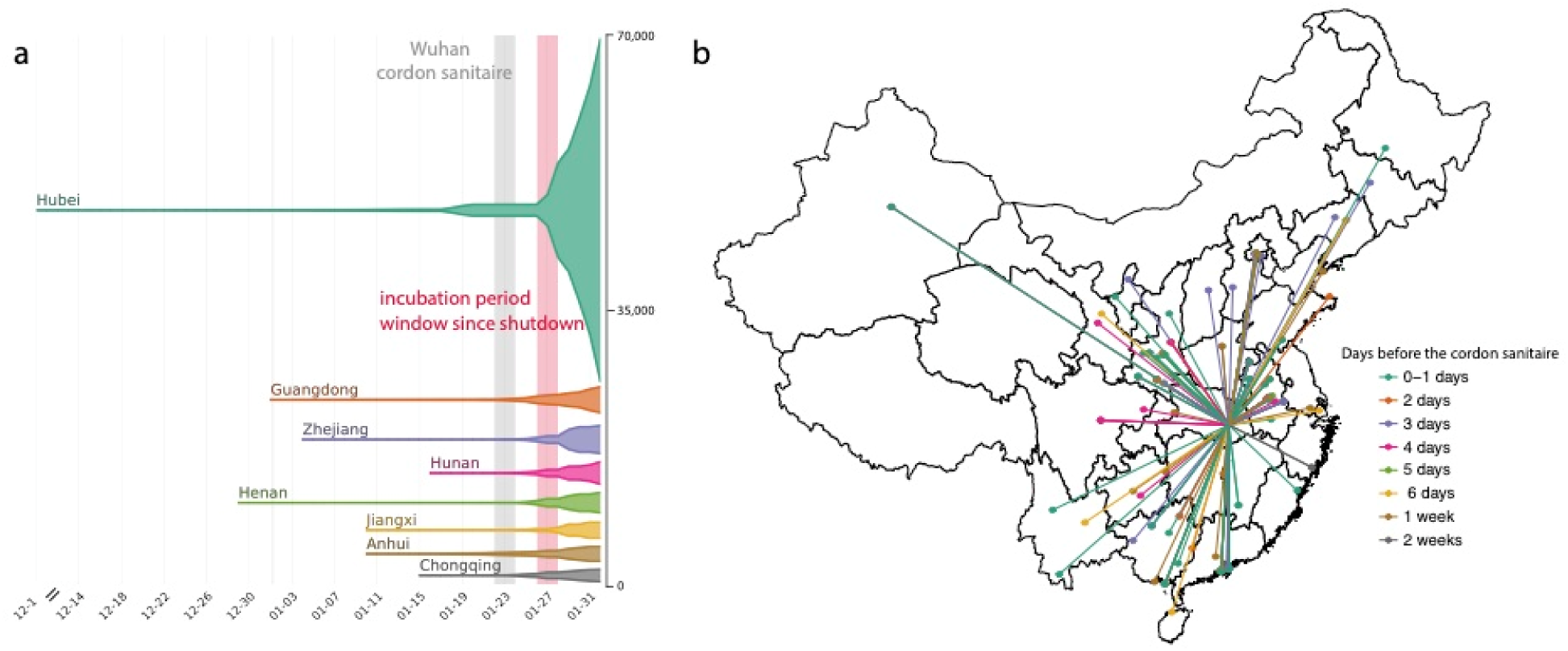
Number of cases and key dates during the epidemic. (a) The epidemic curve of the COVID-19 outbreak in provinces in China. Vertical lines and boxes indicate key dates such as implementation of cordon sanitaire of Wuhan (grey) and the end of the first incubation period after the travel restrictions (red). The thin grey line represents the closure of Wuhan seafood market on 1^st^ January 2020. The width of each horizontal tube represents the number of reported cases in that province. (b) Map of COVID-19 confirmed cases (n = 554) that had reported travel history from Wuhan before travel restrictions were implemented on January 23, 2020. Colors of the arrows indicate date of travel relative to the date of travel restrictions.

To identify accurately a timeframe for evaluating early shifts in SARS-CoV-2 transmission in China, we first estimated from case data the average incubation period of COVID-19 infection (*i*.*e*. the duration between time of infection and symptom onset (*6, 7*)). Since infection events are typically not directly observed, we estimate incubation period from the span of exposure during which infection likely occurred. Using detailed information on 38 cases for whom both the dates of entry to and exit from Wuhan are known, we estimate the mean incubation period to be 5.1 days (std. dev. = 3.0 days; **Fig. S1**), similar to previous estimates from other data (*8, 9*). In subsequent analyses we add an upper estimate of one incubation period (mean + 1 standard deviation = 8 days) to the date of Wuhan shutdown, in order to delineate the date before which cases recorded in other provinces might represent infections acquired in Hubei (i.e. 1^st^ February 2020; **Fig. 1a**).

In order to understand whether the volume of travel within China can predict the epidemic outside of Wuhan, we analyzed real-time human mobility data from Baidu Inc., together with epidemiological data from each province (**Materials and Methods**). Small local transmission clusters (in a family) of COVID-19 were first reported in Guangzhou, Guangdong province (*14*). We investigated spatio-temporal disease spread to elucidate the relative contribution of Wuhan to transmission elsewhere and evaluate how the cordon sanitaire may have impacted it.

Among cases reported outside Hubei province in our dataset, we observe 515 cases with known travel history to Wuhan and a symptom onset date before 31^st^ January 2020, compared with only 39 after 31^st^ January, 2020, illustrating the effect of travel restrictions **(Figs. 1b, 2a, Fig. S3)**. We confirm the expected decline of importation with real-time human mobility data from Baidu Inc. Movements of individuals out of Wuhan increased in the days before the Lunar New Year and the establishment of the cordon sanitaire, before rapidly decreasing to almost no movement **(Figs. 2a,b)**. The travel ban appears to have prevented travel in and out of Wuhan around the time of the Lunar New Year celebration (**Fig. 2a**) and likely reduced further dissemination of SARS-CoV-2 from Wuhan.

**Figure 2:**
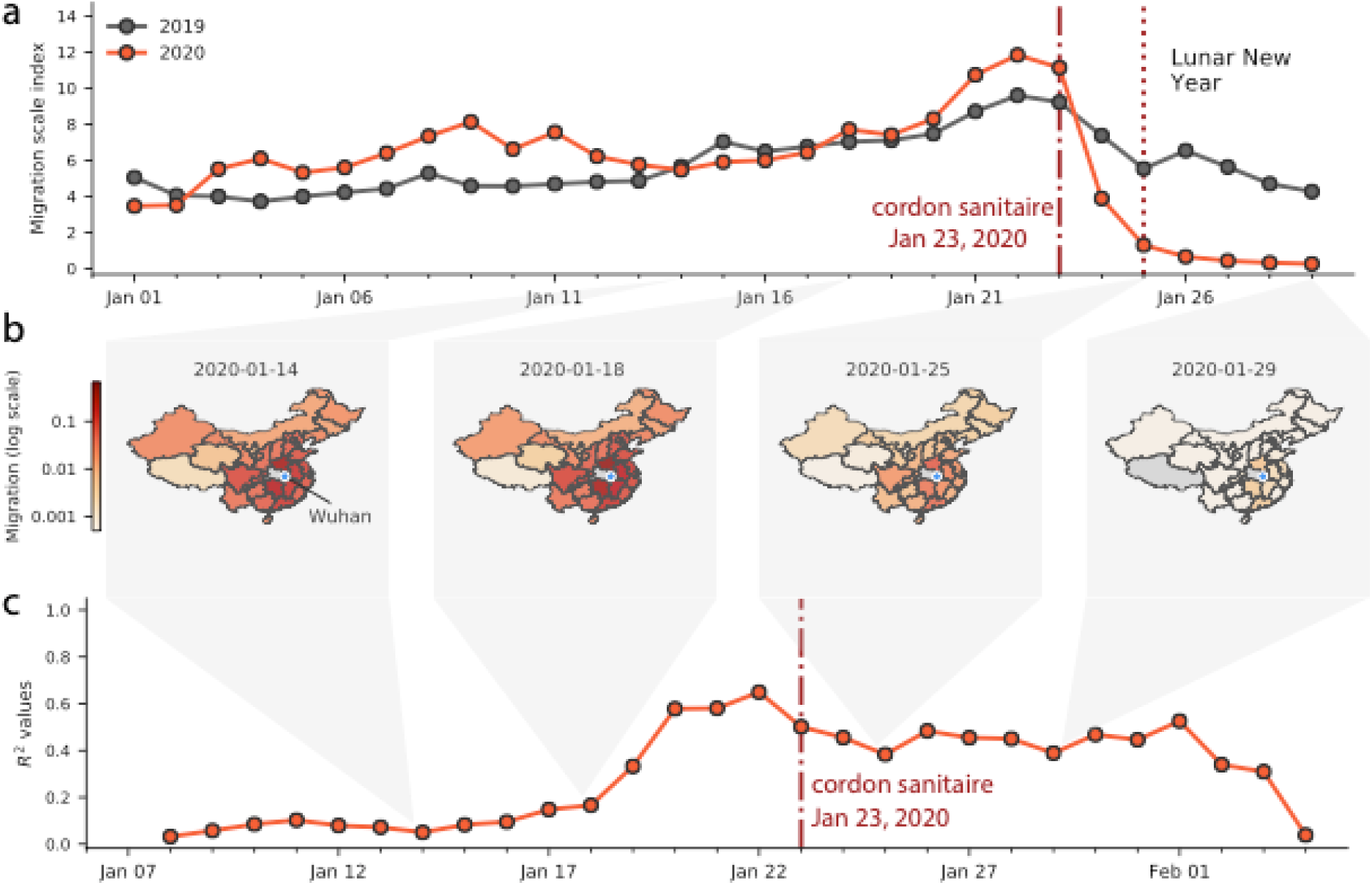
Human mobility, spread and synchrony of COVID-19 outbreak in China. a) Human mobility data extracted in real time from Baidu. Date of start of travel ban out of Wuhan and other control measures on January 23,2020. Dark and red lines represent migration scale indexes for 2019 and 2020, respectively. b) Relative movements from Wuhan to other provinces in China. c) Timeline of daily correlation between daily incidence in Wuhan and incidence in all other provinces weighted by human mobility.

To test the contribution of the epidemic in Wuhan to seeding epidemics elsewhere in China we build a naïve COVID-19 GLM (*15*) model of daily case counts (**Materials and Methods**). We estimate the epidemic doubling time outside Hubei to be 4.0 days (range across provinces of 3.6 - 5.0 days) and estimate the epidemic doubling time within Hubei to be 7.2 days, consistent with previous reports (*5, 9, 16, 17*). Our predicts daily case counts across all provinces with relatively high accuracy (as measured with a pseudo-R^2^ from a negative binomial GLM) throughout early February 2020, and when accounting for human mobility (**Figs. 2c, Table S1, S2**).

We find that the magnitude of the epidemic (total number of cases until February 10, 2020) outside of Wuhan is remarkably well predicted by the volume of human movement out of Wuhan alone (R^2^ = 0.89 from a log-linear regression using cumulative cases, **Fig. S8**). Therefore cases exported from Wuhan prior to the cordon sanitaire appear to have not only contributed to initiating local chains of transmission, both in neighboring provinces such as Henan, and in comparatively more distant provinces, e.g., Guangdong and Zhejiang **(Figs. 1a, 2b)** but that the frequency of introductions from Wuhan are also predictive of the size of the epidemic in other provinces (controlling for population size).

After 1^st^ February 2020 (corresponding to one mean + one SD incubation period after the cordon sanitaire and other interventions were implemented), the correlation of daily case counts and human mobility from Wuhan decreases (**Figs. 2c**) which indicates that variability is better explained by other factors such as local public health response. This suggests that travel restrictions may have reduced the flow of case importations from Wuhan; but also that other factors guiding a local epidemic may be taking over.

We also estimate the growth rates of the epidemic in all other provinces (**Materials and Methods**). Interestingly, we find that all provinces outside Hubei experienced faster growth rates between January 9^th^ – January 22^nd^, 2020 (**Fig. 3b, Fig. S4b**) which was the time before travel restrictions and drastic control measures were implemented (**Fig. 3c, Fig. S6**); this is also apparent from the case counts by province (**Fig. S6**). In the same period, we show that variation in the growth rates are almost entirely explained by human movements from Wuhan **(Fig. 3c**), consistent with theory of infectious disease spread in highly coupled metapopulations (*19, 20*). Following the implementation of drastic control measures across the country, growth rates become negative (Fig 3b), indicating that transmission is being successfully mitigated. The correlation of growth rates and human mobility from Wuhan becomes negative, i.e. provinces with larger mobility prior to the cordon sanitaire (but also larger number of cases overall) have more rapidly declining growth rates of daily case counts. This could be partly due to travel restrictions but also the fact that control measures may have been more drastic in locations with larger outbreaks with local transmission (see more detail in section **“Current role of imported cases in Chinese provinces”)**.

**Figure 3:**
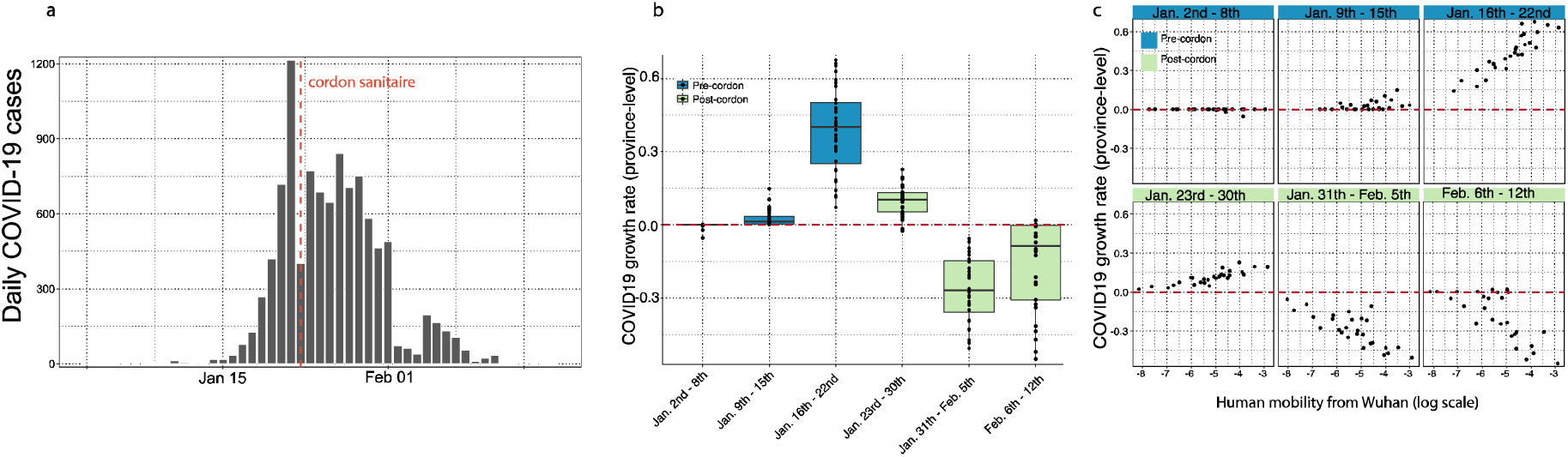
Human mobility explains growth rate of epidemic in China. (a) Daily case counts of cases in China. (b) Time series of province-level growth rates of the COVID-19 epidemic in China. Estimates of the growth rate were obtained by performing a time-series analysis using mixed-effect model of lagged, log linear daily case counts in each province (**Materials and Methods**). Above the red line are positive growth rates and below are growth rates that are negative. Blue indicates before the implementation of the cordon sanitaire and green after. c) Relationship between the growth rate and human mobility at different times of the epidemic. Blue indicates before the implementation of the cordon sanitaire and green after.

The travel ban coincides with increased testing capacity across provinces in China. An alternative hypothesis is that the observed epidemiological patterns outside Wuhan are the result of increased testing capacity. We test this hypothesis by including differences in testing capacity before and after the rollout of large scale testing in China on 20^th^ of January 2020 and test its impact on the predictability of daily cases (**Materials and Methods**). We plot the relative improvement in the prediction of our model (based on normalized residual error) of (i) a model that includes mobility from Wuhan and (ii) a model that includes testing availability (see more details in **Materials and Methods**). Overall, the inclusion of mobility data from Wuhan produces a significant improvement in the models prediction (delta-BIC > 250, (*24*)) over a naive model that considers only autochthonous transmission with a doubling time of 2-8 days **(Fig. 3b)**. Of the 27 provinces in China reporting cases through February 6^th^, 2020, we find that in 12 provinces the largest improvements in prediction can be achieved using mobility only (**Fig. S5**). In 10 provinces, both testing and mobility improve the models prediction and in only one province testing is the most important factor improving model prediction (**Fig. S5**). We conclude that laboratory testing during the early phase of the epidemic is critical, however, mobility out of Wuhan remains the main driver of spread prior to the shutdown.

### Current role of imported cases in Chinese provinces

As case counts are decreasing outside Wuhan (Fig 3b), we can further investigate the current contribution of imported cases to local epidemics outside Wuhan by investigating case characteristics. Age and sex distributions can reflect heterogeneities in the risk of infection within affected populations. To investigate meaningful shifts in the epidemiology of the COVID-19 outbreak through time, we examined age and sex data for cases from different periods of the outbreak, and from individuals with and without travel from Wuhan. However, details of travel history exist for only a fraction of confirmed cases and this information is particularly scant for some provinces (e.g. Zhejiang and Guangdong). Consequently, we grouped confirmed cases into four categories: (*I*) early cases with travel history (early = reported before 1^st^ Feb), (*II*) early cases without travel history, (*III*) later cases with travel history (later = reported between 1^st^ – 10^th^ Feb), (*IV*) later cases without travel history.

Using crowdsourced case data, we found that cases with travel history (categories *I* and *III*) had similar median ages and sex ratios in both the early and later phases of the outbreak (41 vs 42 years old, 50% interquartile interval: 32.75 vs 30.75 and 54.25 vs 53.5 respectively, p-value > 0.1; 1.47 vs. 1.45 males per female, respectively; **Fig. 4d, Fig. S7**). Early cases with no information on travel history (category *II)* had a similar median age and sex ratio to those with known travel history (42 years old (50% interquartile interval: 30.5 – 49.5, p-value > 0.1) and 1.80 males per female, **Fig. 4d**). However, the sex ratio of later cases without reported travel history (category *IV*) shifted to approximately 1:1 (57 male vs. 62 female, X_2_ test, p-value < 0.01), as expected under a null hypothesis of equal transmission risk **(Fig. 4a**,**b**,**d**, see also reference (*11, 12*), **Materials and Methods)** and the median age in this group increased to 46 (50% interquartile interval: 34.25 – 58, t-test: p-value < 0.01) (**Figs. 4a**,**b**,**c, Fig. S7**). We hypothesize that many of the cases with no known travel history in the early period were indeed travelers that contributed to disseminating SARS-CoV-2 outside of Wuhan. The shift towards more equal sex ratios and older ages in non-travellers after 31^st^ January 2020 confirm the finding that epidemics outside Wuhan were then driven by local transmission dynamics. The case definition changed to include cases without travel history to Wuhan after 23^th^ January 2020 (**Materials and Methods**).

**Figure 4:**
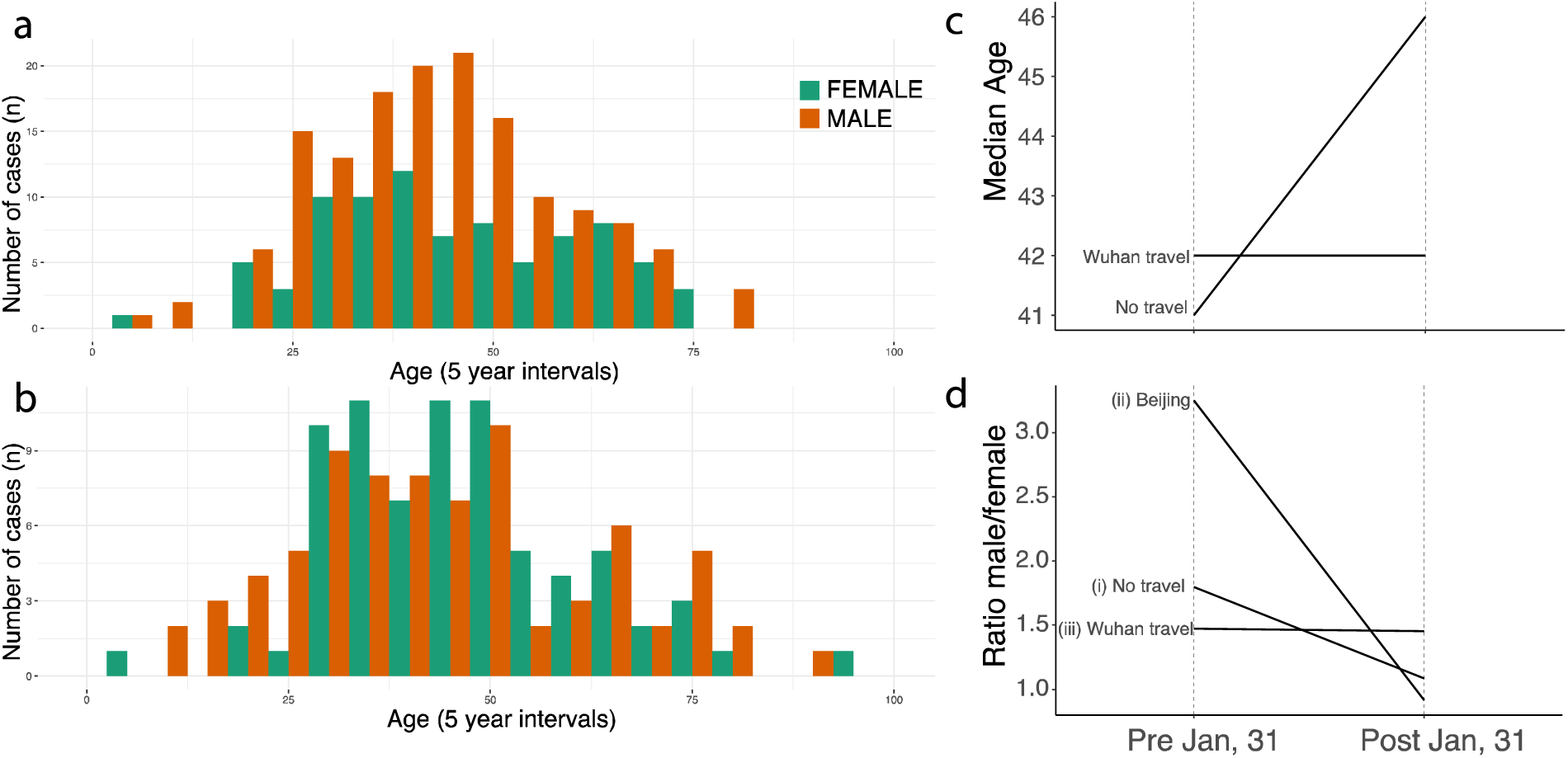
Shifting age and sex distributions through time. (a) Age and sex distributions of confirmed cases with known travel history to Wuhan, (b) Age and sex distributions of confirmed cases that had no travel history. (c) Median age for cases reported early (before 1^st^ Feb) and those reported later (between 1^st^ – 10^th^ February. Uncertainty is shown in Fig. S7. (d) Change through time in the sex ratio of (i) all reported cases in China with no reported travel history, (ii) cases reported in Beijing without travel history, and (iii) cases known to have travelled from Wuhan.

## Discussion

Containment of respiratory infections is particularly difficult if they are characterized by relatively mild symptoms or transmission before the onset of disease (*25*). Intensive control measures, including travel restrictions, have been implemented to limit the spread of COVID-19 in China. Here, we showed that this combination of interventions was successful in mitigating the spread and reduce local transmission of COVID-19 in China although it is not possible to definitively determine the individual impact of each intervention. More analyses will be required to determine how to optimally balance the expected positive effects on health with the negative impact on individual liberties, the economy, and society at large.

## Data Availability

Code and data are available on the following GitHub repository: https://github.com/Emergent-Epidemics/covid19_cordon

## Acknowledgements

We would like to thank all individuals who are collecting epidemiological data of the COVID-19 outbreak around the world.

## Authors contributions

MUGK, OGP, SVS developed the idea and research. MUGK and SVS wrote the first draft of the manuscript and all other authors discussed results and edited the manuscript. MUGK, BG, SVS, DMP and the open COVID-19 data working group collected and validated epidemiological data. RL and MUGK collected intervention data. C-HY, BK, and SVS collected and processed human mobility data.

## Materials and Methods

### Epidemiological data

No officially reported line list was available for cases in China (*26*). We use a standard protocol (*27, 28*) to extract individual level data from December 1st, 2019 - February 10^th^, 2020. Sources are mainly official reports from provincial, municipal or national health governments. Data included basic demographics (age, sex), travel histories and key dates (dates of onset of symptoms, hospitalization, and confirmation). Data were entered by a team of data curators on a rolling basis and technical validation and geo-positioning protocols were applied continuously to ensure validity. A detailed description of the methodology is available (*27*) (attached in **Supplementary information**). Lastly, total numbers were matched with officially reported data from China and other government reports. For sensitivity, GLM analyses (see below) were performed with case counts from the World Health Organization.

### Proportions of symptomatic travelers

The proportion of cases who travelled while symptomatic was assessed from a subset of 236 cases for whom the dates of symptom onset and departure from Wuhan were available. Residency was split into three categories: Wuhan, China and International. Foreigners living in Wuhan were categorized as Wuhan and patients with missing Wuhan residency were either kept as missing values or categorized according to their country of origin. Both parametric (χ^2^ test, (*29*)) and non-parametric (exact Fisher test, (*30*)) tests were performed and the uncertainty in proportions was assessed by the standard deviation of sample proportions.

### Statistical inference of the incubation period

The incubation period is the time interval between infection and symptom onset. We assumed that cases travelling from Wuhan were exposed during their stay in Wuhan. We estimated the incubation period from 38 travelling cases returning from Wuhan with known dates of symptom onset, entry and exit. The end of the exposure period was assumed to be the exit travel date except if symptom onset occurred prior to the exit date (in which case exposure was assumed to have occurred prior to symptom onset). The start of the exposure period corresponded to the entry date. We assumed that the incubation period could not exceed 30 days.

For each case, the minimum and maximum incubation period was derived from the dates of entry, exit and symptom onset

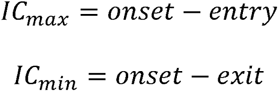

We fitted a truncated gamma distribution (0 to 30 days) and estimated the mean and variance of the incubation period using Markov Chain Monte Carlo (MCMC) in a Bayesian framework using an uninformative prior distribution. We derived the likelihood as follows:

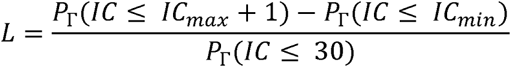

A Metropolis-Hastings algorithm was implemented in R. Marginal posteriors were sampled from a chain of 5,000 steps after discarding a burn-in of 50 steps. Convergence was inspected visually.

### Models of shifting age and sex distributions

Age and sex distributions are important in understanding risk of infection across populations. Assuming risk to be distributed relatively equally across a population, as an outbreak evolves age and sex distributions should follow the underlying population structure. Varying degrees of immunity and exposure may shift these distributions (*31*). To examine whether the ongoing outbreak shifted from an epidemic concentrated in Wuhan and among travelers from Wuhan to an epidemic that was self-sustained in provinces across China we use age and sex data from different periods of the outbreak for individuals with reported travel history and no known travel history. We define two periods of the outbreak, an “early” phase, starting with the first reports in early December and ending a set number of days after the Wuhan shutdown. This was selected to be 8 days after the Wuhan shutdown, which conservatively corresponds to one incubation period + 1SD (see above) after the shutdown. After that date (i.e. 1^st^ Feb 2020) we assume that most reported transmissions in provinces outside of Wuhan are the result of local transmission. We further divided our data in those that had cases with known travel history to Wuhan and those who did not. Then we produce the following summary statistics:

1 Average age stratified by sex for all cases with reported travel history to Wuhan.
2 Average age stratified by sex for all cases with no reported travel history to Wuhan in the period between December 1, 2019 - January 31, 2020.

We then compare these with:

3 Average age stratified by sex for all cases with no reported travel history to Wuhan in the period between January 31, 2020 - February 10, 2020.

*Model M1* compares the distribution of age and sex among travelers to the reported infections outside Wuhan with no known travel history. In case these two distributions are similar, import driven epidemic can be concluded. Under our *model assumptions M2*, if the epidemic was driven largely by importations across the two time periods, all age and sex distributions should mirror those of the reported traveler infections. Under our *model* assumption *M3*, if the epidemic was driven by other factors (i.e., local transmission), the two distributions should vary across the two time periods.

We cannot exclude the possibility that shifts in distributions may be due to heightened awareness among the general population which may have increased reporting in female cases later in the epidemic. Further, more work will be necessary to understand the differential risk of severe or symptomatic disease to fully understand the age and sex distributions in this outbreak. For example, why there are relatively few reports of cases <18y old. However, as for other respiratory pathogens symptomatic and severe infection were more concentrated in older populations. We do not intend to make any general statements about differential risk but were more interested in shifts in reported cases across multiple geographies in China.

### Real time human mobility data

We extract human mobility data from the Baidu Qianxi web platform, which presents daily population travels between cities or provinces tracked through the Baidu Huiyan system. The data do not represent numbers of individual travelers but rather an index of relative movements constructed by Baidu’s proprietary methods which are correlated with human mobility (*32*) (http://qianxi.baidu.com/). In particular, two pieces of information are collected. First, we extract a series of migration scale indices for traveling out of Wuhan, from January 1st to February 10, both in 2019 and 2020. Second, we obtain the proportion of human movement from Wuhan were bound for each of 31 provinces in China. These proportions are available for January 1st - February 10, 2020. Based on this data we had access to both changes in mobility volume and changes in mobility direction. See more detailed descriptions of the human movement data here: (*33, 34*)

### Review of interventions and reporting shifts

We reviewed the literature and online social media to understand the key timings of interventions and announcements that are relevant for disease transmission across China. We collated information about the type (e.g., announcement of outbreak, travel restrictions, isolation of patients, etc.), geographic location (e.g., city where available, province), and timing (specific date or date range).

### COVID-19 case definitions

Definitions of probable and confirmed COVID-19 cases have changed throughout the epidemic. We collected data from official sources describing the timing and specifics of the case definitions.

*From January 18-22:*

**Probable:** Need to satisfy (i) and (ii):

i Clinical symptoms: (1) fever; (2) imaging showing pneumonia typical of the disease; (3) during early disease, total white cells normal or reduced, or lymph cell count reduced.
ii Epidemiologic history: (1) within 2 weeks of symptom onset, Wuhan travel or resident history; or within 2 weeks of symptom onset, contact with persons from Wuhan who had fever with respiratory symptoms; or belong to a cluster.

**Confirmed**: Need to satisfy criteria for probable case and have a real-time quantitative polymerase chain reaction (RT-qPCR) positive result from sputum, nasopharyngeal swabs, lower respiratory tract secretions or other sample tissue, or genome sequencing highly similar with known SARS-CoV-2. available strains.

*From January 22-23:*

**Probable**: Need to satisfy (i) and any one epidemiologic history described in (ii):

i Clinical symptoms: (1) fever; (2) imaging showing pneumonia typical of the disease; (3) during early disease, total white cells normal or reduced, or lymph cell count reduced
ii Epidemiologic history: (1) within 2 weeks of symptom onset, Wuhan travel or resident history; (2) within 2 weeks of symptom onset, contact with persons from Wuhan who had fever with respiratory symptoms; (3) belong to a cluster or had epidemiologic link with confirmed cases.

**Confirmed:** Need to satisfy criteria for probable case and have a RT-qPCR positive result from respiratory or blood samples, or genome sequencing highly similar with known SARS-CoV-2. available strains.

*From January 23-27:*

**Probable:** Need to satisfy (i) and (ii):

i Clinical symptoms: (1) fever; (2) imaging showing pneumonia typical of the disease; (3) during early disease, total white cells normal or reduced, or lymph cell count reduced
ii Epidemiologic history: within 2 weeks of symptom onset, Wuhan travel or resident history; or within 2 weeks of symptom onset, contact with persons from Wuhan who had fever with respiratory symptoms, or belong to a cluster.

**Confirmed:** Need to satisfy criteria for probable case and have a RT-qPCR positive result from sputum, nasopharyngeal swabs, lower respiratory tract secretions, or other samples, or genome sequencing highly similar with known SARS-CoV-2. available strains.

*From January 27-February 5:*

**Probable:** Need to satisfy any two of the symptoms described in (i) and any of the epidemiological history described in (ii):

i Clinical symptoms: (1) fever; (2) imaging showing pneumonia typical of the disease; (3) during early disease, total white cells normal or reduced, or lymph cell count reduced
ii Epidemiologic history: (1) within 2 weeks of symptom onset, travel or resident history in Wuhan region or other places with sustained local transmission; (2) within 2 weeks of symptom onset, contact with persons from Wuhan city or other places with sustained local transmission who had fever with respiratory symptoms, (3) belong to a cluster or epidemiologic connection with COVID-19 infected persons.

**Confirmed**: Need to satisfy criteria for probable case and have a RT-qPCR positive result from respiratory or blood samples, or genome sequencing highly similar with known SARS-CoV-2. available strains from lab test of respiratory or blood samples.

### Comparing predictive models of epidemic trajectories

To evaluate hypotheses regarding the effect of mobility and testing on COVID-19 dynamics, we fit three different Generalized Linear Models (GLM). Model 1 was a Poisson GLM to estimate daily case counts, Model 2 was a negative binomial GLM to estimate daily case counts, and Model 3 was a log-linear regression to estimate daily cumulative cases. BIC scores shown in Fig. 4b are calculated on a GLM of the form Y(t) = Y(t-4) + IT(t) + M(t-5) + IM(t) where Y(t) is either the number of new cases observed on day t (Model 1 & 2) or cumulative number of cases observed through day t (Model 3), Y(t-4) represents the number of cases (or the cumulative number under Model 3) four days prior (median doubling time outside Hubei province), IT(t) is an indicator function for PCR test availability that is 1 after 19th January 2020 and 0 before, M(t-5) is the Baidu.Inc-estimated mobility between Wuhan and each province 5 days prior (median incubation period), and IM(t) is an indicator function which is set to 1 after 26th January 2020 and 0 before (which represents one median incubation period from 22nd January 2020). Models were fit to province-level data. The three models were compared using differences in Bayesian Information Criteria (BIC), where larger values indicate models with lower relative support, and BIC>4 considered the cutoff for substantial model improvement. We performed a detailed sensitivity analysis on the availability of PCR tests, doubling time, and incubation periods. We obtained qualitatively similar results for Model 1 (Poisson GLM fit to daily case counts), Model 2 (negative binomial GLM fit to daily case counts), and Model 3 (log-linear regressions fit to cumulative cases), see Table S2. In addition, we provide a full time series analysis of the optimal lag structure for cases and mobility for each province. Additionally, although BIC is considered more conservative, model selection results were confirmed using AIC for model selection (see Fig. 4 and Table S2). Lastly, we validated our model selection results using elastic-net regression and n-fold cross validation as implemented in the R package GLMNET (*11, 12*.

### Estimating epidemic doubling time

To estimate the epidemic doubling time across each province, we fit a mixed effects Poisson GLM of daily case counts to days since the first case report in each province (fixed effect) and a random effect for each province on the slope and intercept, using the R package lme4 v.1.1-21 (*35*). Daily case counts were determined using the date of symptom onset. However, we only have a symptom onset date for 667 cases. Where the date of symptom onset was not available, we estimated the symptom onset date based on a linear regression model where symptom onset date was fit to confirmation date (n =632 with both onset and confirmation dates, p < 0.001, R^2^ = 0.77). Using this model, we estimated the onset date for the 31436 cases with a recorded confirmation date.

### Model selection via elastic-net regression and cross-validation

We fit regularized Poisson and negative binomial models with an elastic net penalty, i.e., 50/50 mixture of the lasso and ridge penalties, with the regularization coefficient (lambda) selected by leave-one-out cross-validation. As seen with the AIC/BIC-based model selection, the regularized model included terms for lagged cases, the mobility and testing indicator variables, and mobility out of Wuhan. The out-of-sample log likelihood for the regularized Poisson regression was -9102 and was -22519 for the negative binomial model. The significantly worse fit for the negative binomial model was primarily driven by two outlier predictions, removing those results in an out-of-sample log likelihood for the negative binomial model of -11625.

Models were implemented in the R package glmnet v. 2.0-18 (*36*). Because glmnet has not implemented a negative binomial model, we performed regularization using the Poisson model and estimated the overdispersion parameter using the glm.nb function in the R package MASS v. 7.3-51.4 (*37*).

### Supplementary text

To ascertain whether earlier travel restrictions could have prevented the wide-spread increase in cases witnessed in late-January we constructed a simple forecasting model for COVID-19. Briefly, we forecast the cumulative number of cases in each Chinese province by simply doubling the number of cumulative cases reported six days prior. For dates prior to Jan. 28th and after Feb 3rd, this naive forecast produces an accurate estimate of the cumulative number of cases in each province **(Fig. S4)**. However, the cumulative number of cases reported on Jan 28th is poorly estimated using this model (**Fig. S4)**. In order to accurately forecast the number of cases on Jan 28th, we must also include the relative amount of mobility out of Wuhan into various provinces in the regression model. In **Fig. S4**, we show how a model including only movement from Wuhan on January 22nd fit to the residuals from **Fig. S4** is once again able to accurately forecast cumulative cases. This indicates that for any hope of success, movement restrictions must be prompt.

## Supplementary Tables

**Table S1:**
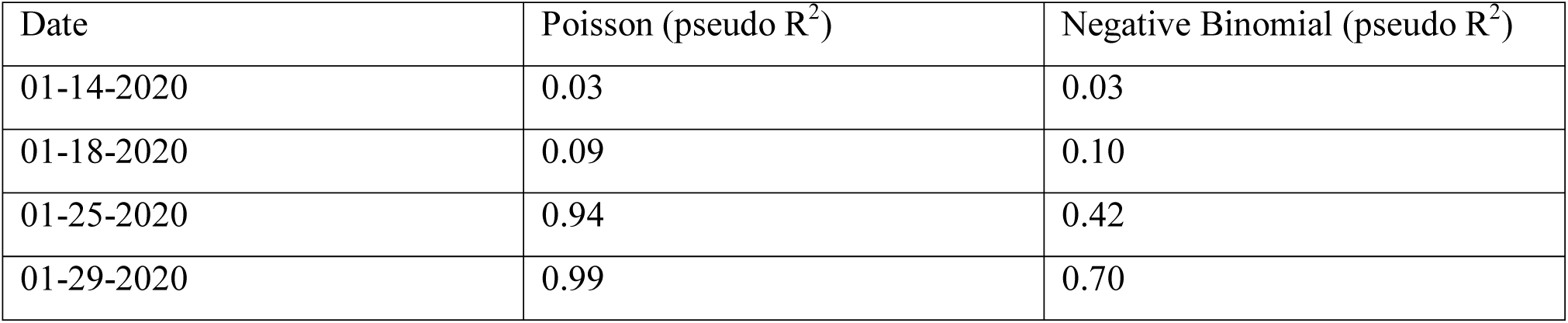
Table shows the pseudo-R^2^ values for Poisson and Negative Binomial GLM of daily case counts and 5-day lagged log mobility from Wuhan, where pseudo-R^2^ were calculated using model deviances as described (*38, 39*).

**Table S2:** Table shows the AIC and BIC values for a log-linear regression based on cumulative cases, a Poisson GLM of daily case counts and a Negative Binomial GLM of daily case counts using seven combinations of predictors.

| Model | LM-AIC | Pois-AIC | NB-AIC | LM-BIC | Pois-BIC | NB-BIC |
| --- | --- | --- | --- | --- | --- | --- |
| CASES_lag4 | 6339.0982<br>6 | 38354.086 | 6805.2375<br>4 | 6356.4876<br>7 | 38365.678<br>9 | 6822.6269<br>4 |
| CASES_lag4-TEST | 6239.9357<br>5 | 28208.904<br>6 | 6482.5221<br>4 | 6263.1216<br>2 | 28226.294 | 6505.7080<br>2 |
| CASES_lag4-MOB | 5538.2643<br>6 | 31090.651<br>3 | 6134.4145<br>8 | 5560.7706<br>4 | 31107.531 | 6156.9208<br>6 |
| CASES_lag4-MOB_IND | 6310.4825<br>4 | 38191.602<br>8 | 6807.2156<br>3 | 6333.6684<br>1 | 38208.992<br>2 | 6830.4015 |
| CASES_lag4-TEST-MOB | 5405.2815<br>6 | 21068.259<br>5 | 5729.1767<br>7 | 5433.4144<br>1 | 21090.765<br>8 | 5757.3096<br>2 |
| CASES_lag4-MOB_IND-MOB | 5520.7081<br>5 | 31000.071 | 6133.9615<br>6 | 5548.841 | 31022.577<br>3 | 6162.0944<br>2 |
| CASES_lag4-MOB_IND-MOB-TEST | 4971.6195<br>4 | 17807.457<br>7 | 5676.4389<br>1 | 5005.3789<br>6 | 17835.590<br>6 | 5710.1983<br>3 |

## Supplementary Figures

**Figure S1:**
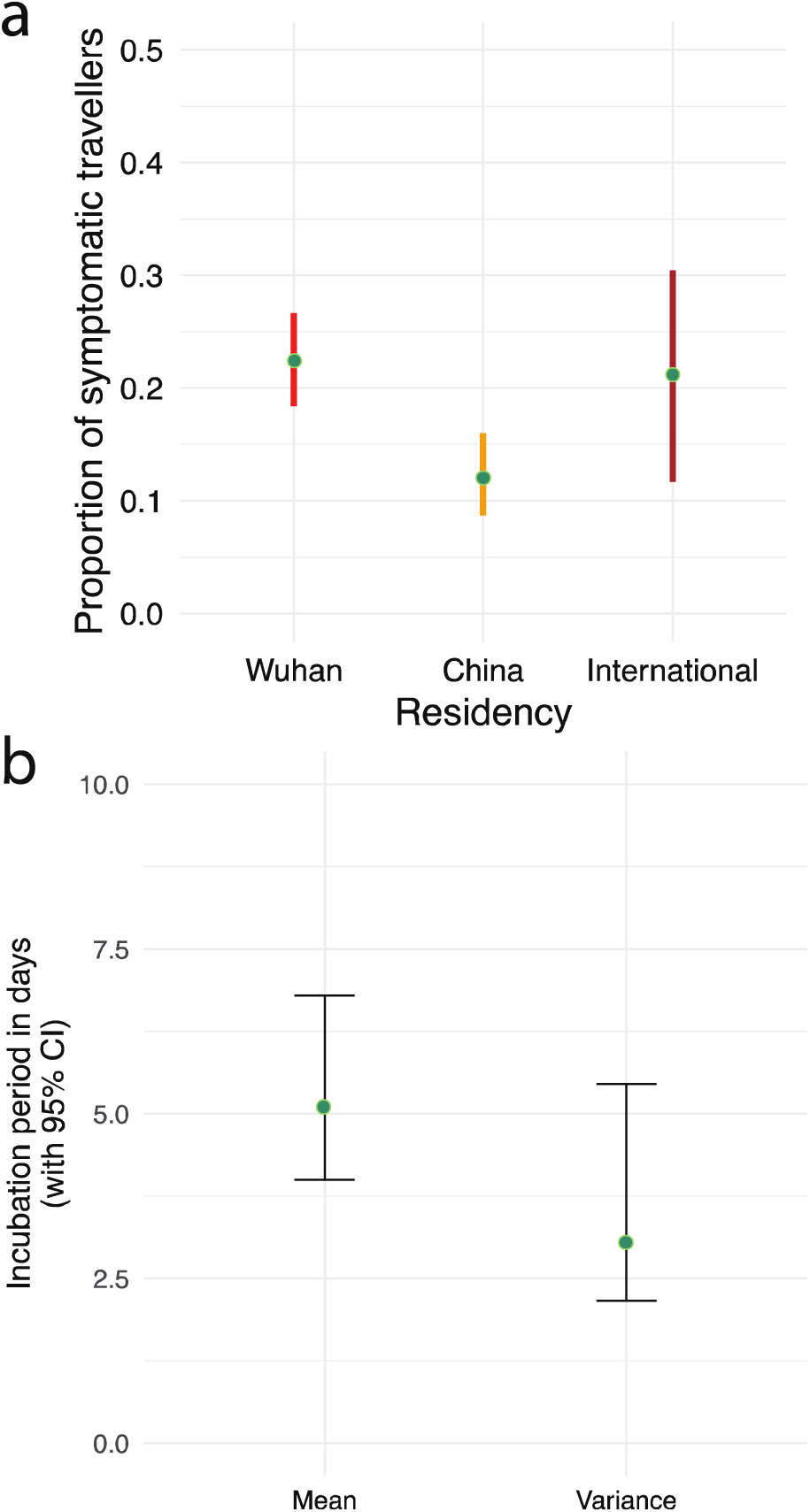
a) Dates of symptom onset before date of travel from Wuhan. b) Incubation period estimates and standard deviation.

**Figure S2:**
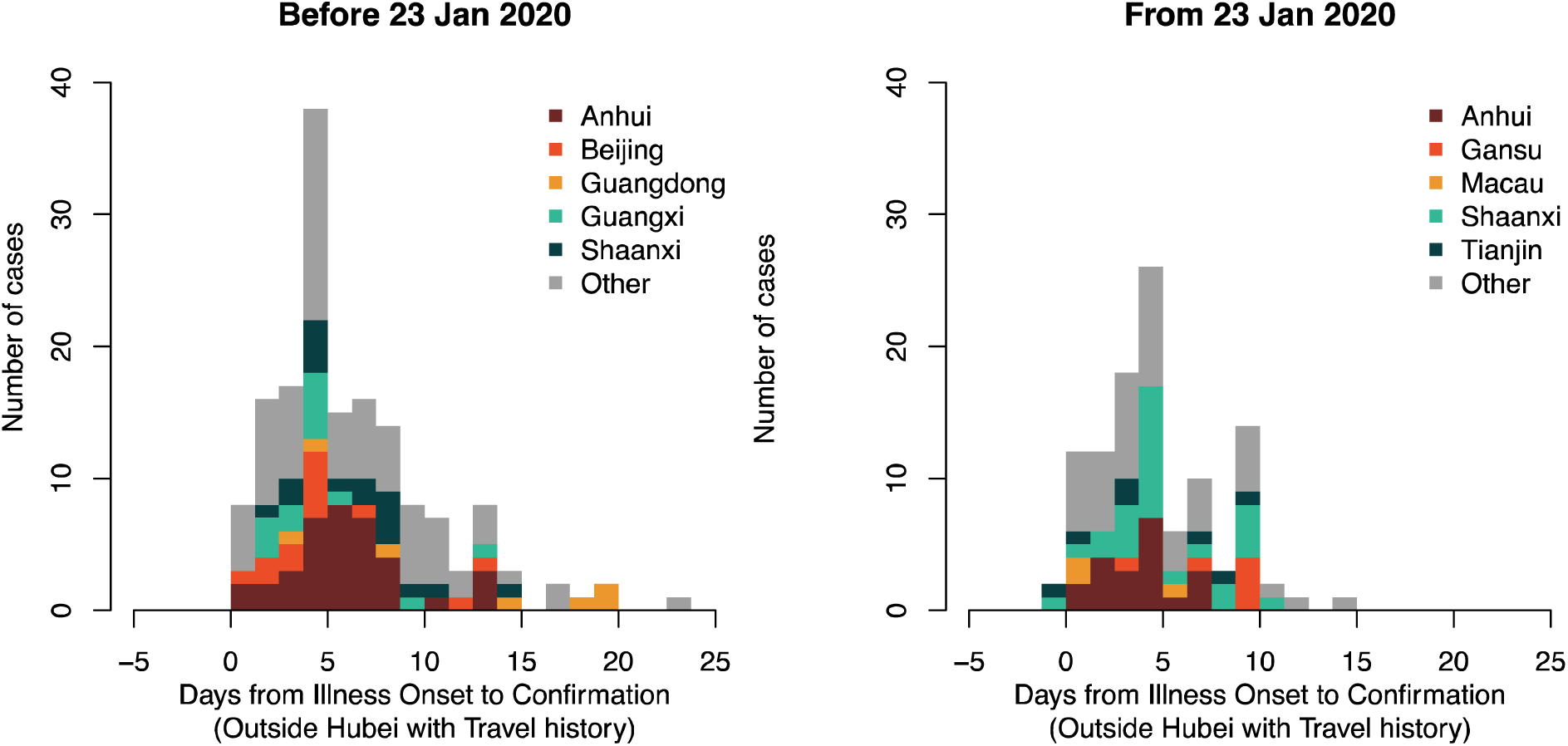
Interval between symptom onset and date of confirmation in confirmed cases with reported travel history in two key periods, before and after January 23, 2020.

**Figure S3:**
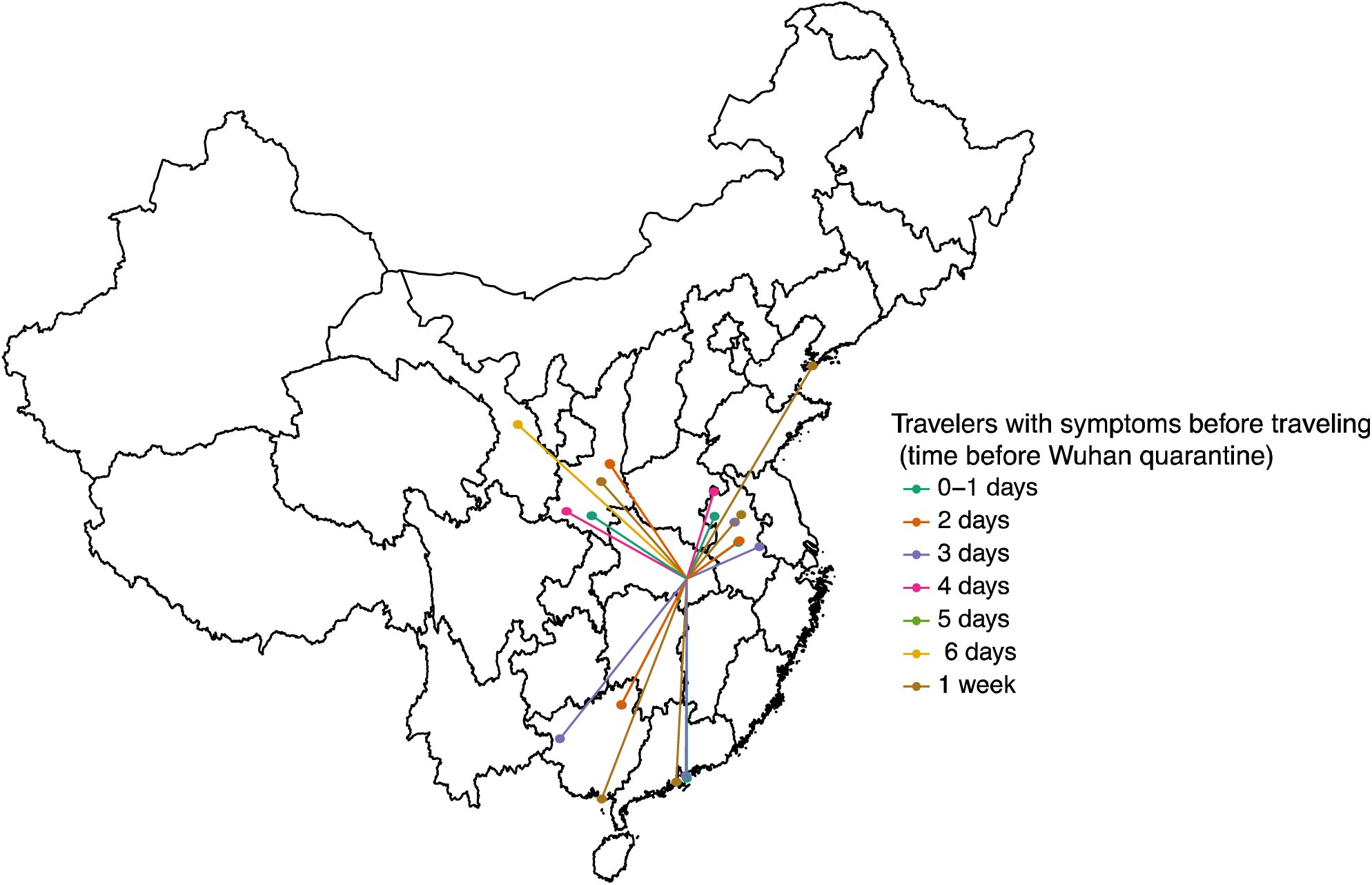
Map of confirmed cases of COVID-19 with known travel history and date of onset date before date of travel.

**Figure S4:**
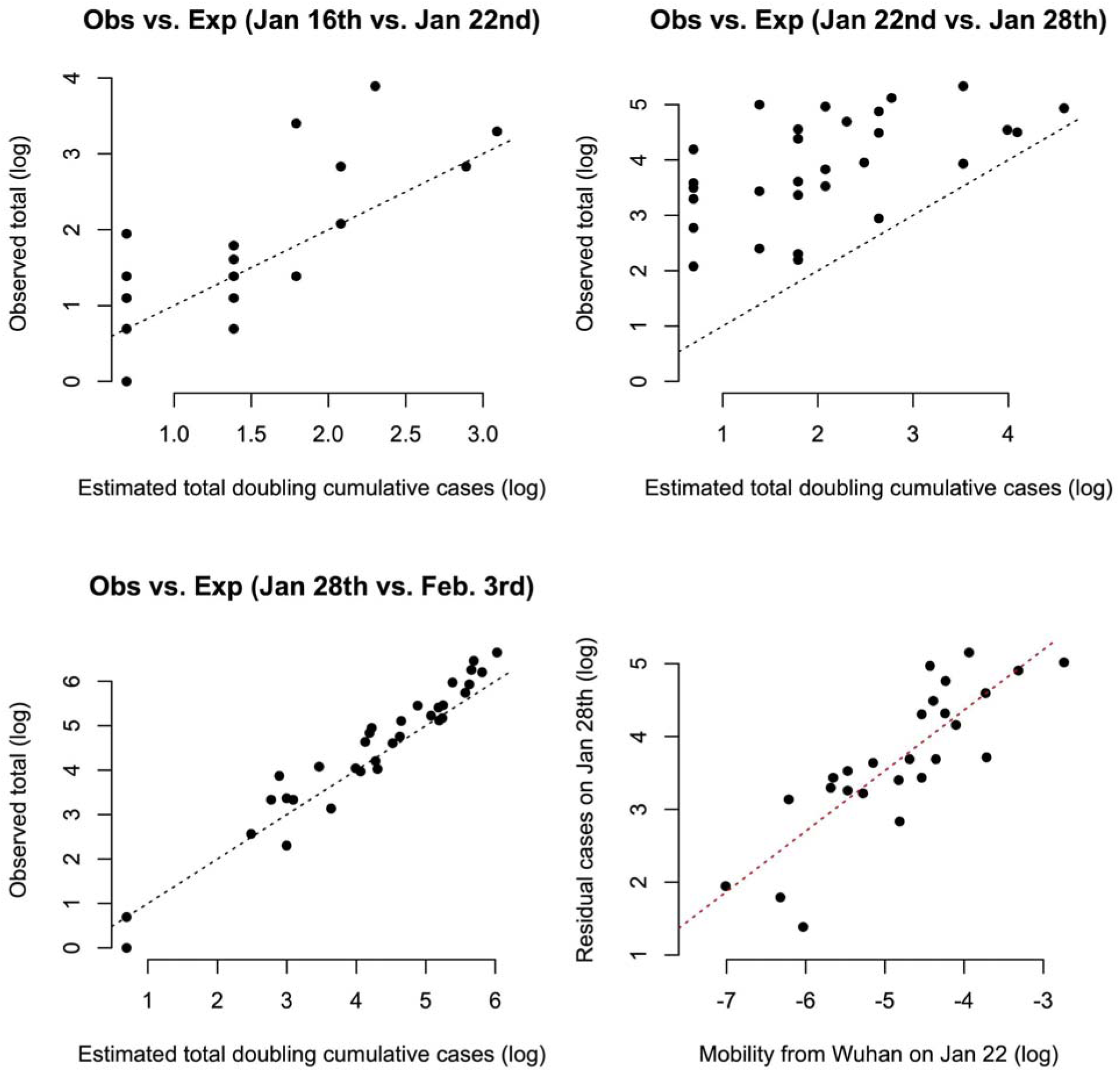
Predicting COVID-19 cases using mobility data. a) Province-level cumulative cases on January 22nd can be accurately predicted based on simply doubling the number of cumulative number of cases occurring on January 16th. b) However, by Jan. 28th, the expected number of cases has significantly increased with respect to predictions based on cases through January 22nd. c) By Feb. 34rd, cumulative cases are once again well estimated based on the cumulative number of cases in each province six days earlier, i.e., on Jan. 28th. d) The deviation in cases on January 28th is well explained by the relative amount of migration out of Wuhan on January 22nd.

**Figure S5:**
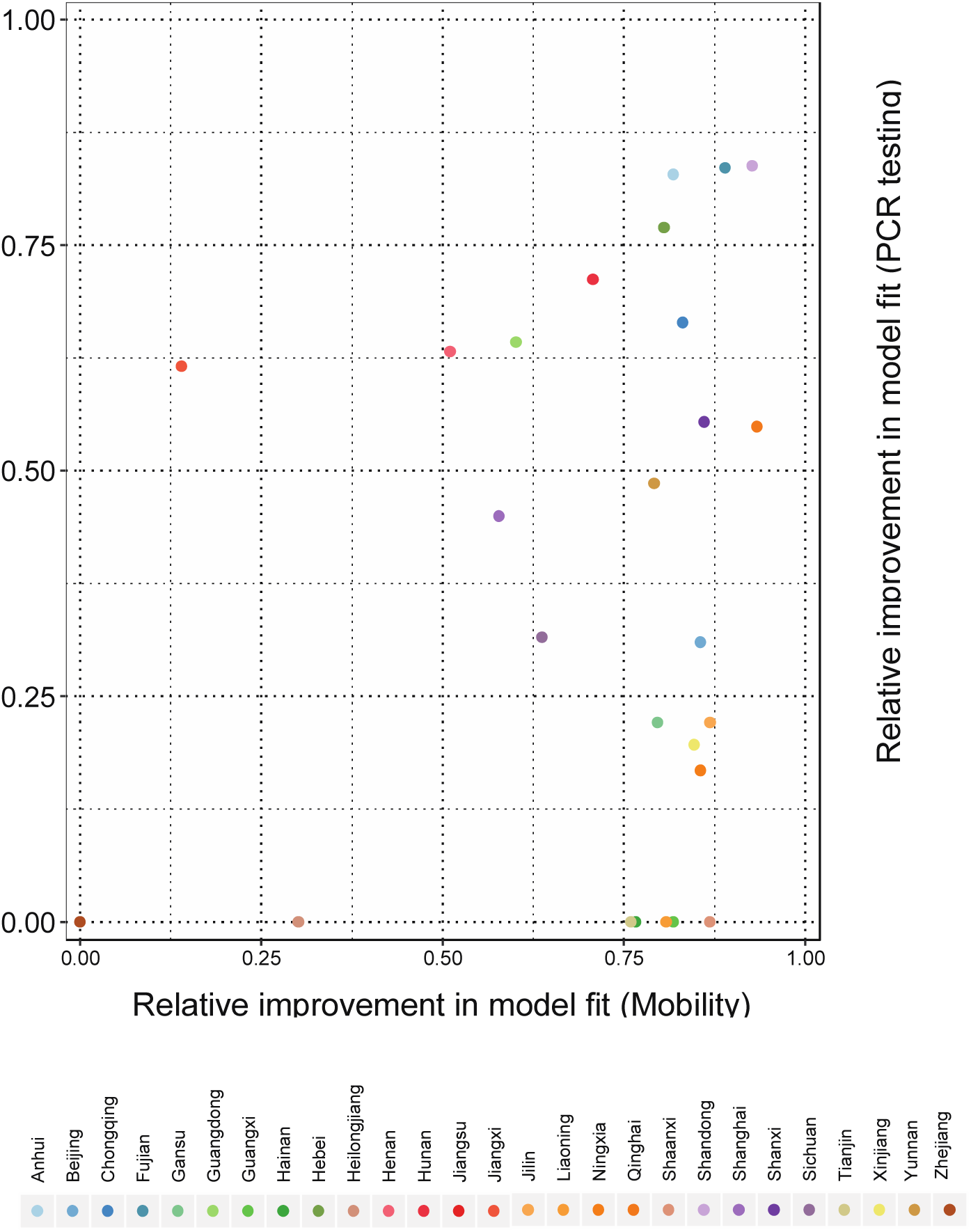
Relative importance of PCR testing vs. human mobility to improve a simple GLM of COVID-19 when estimating exponential growth in province-level cases. Relative improvement is measured as one minus the residuals of a GLM with lagged cases + PCR testing availability (y-axis) and a GLM with lagged cases + mobility from Wuhan. Values were normalized by the observed number of cases such that they ranged between 0 and 1. The resulting metric has a value of 0 for a model where the residual error vastly eclipses the observed data and a value of 1 when residual error is 0, i.e., a perfect model fit.

**Figure S6:**
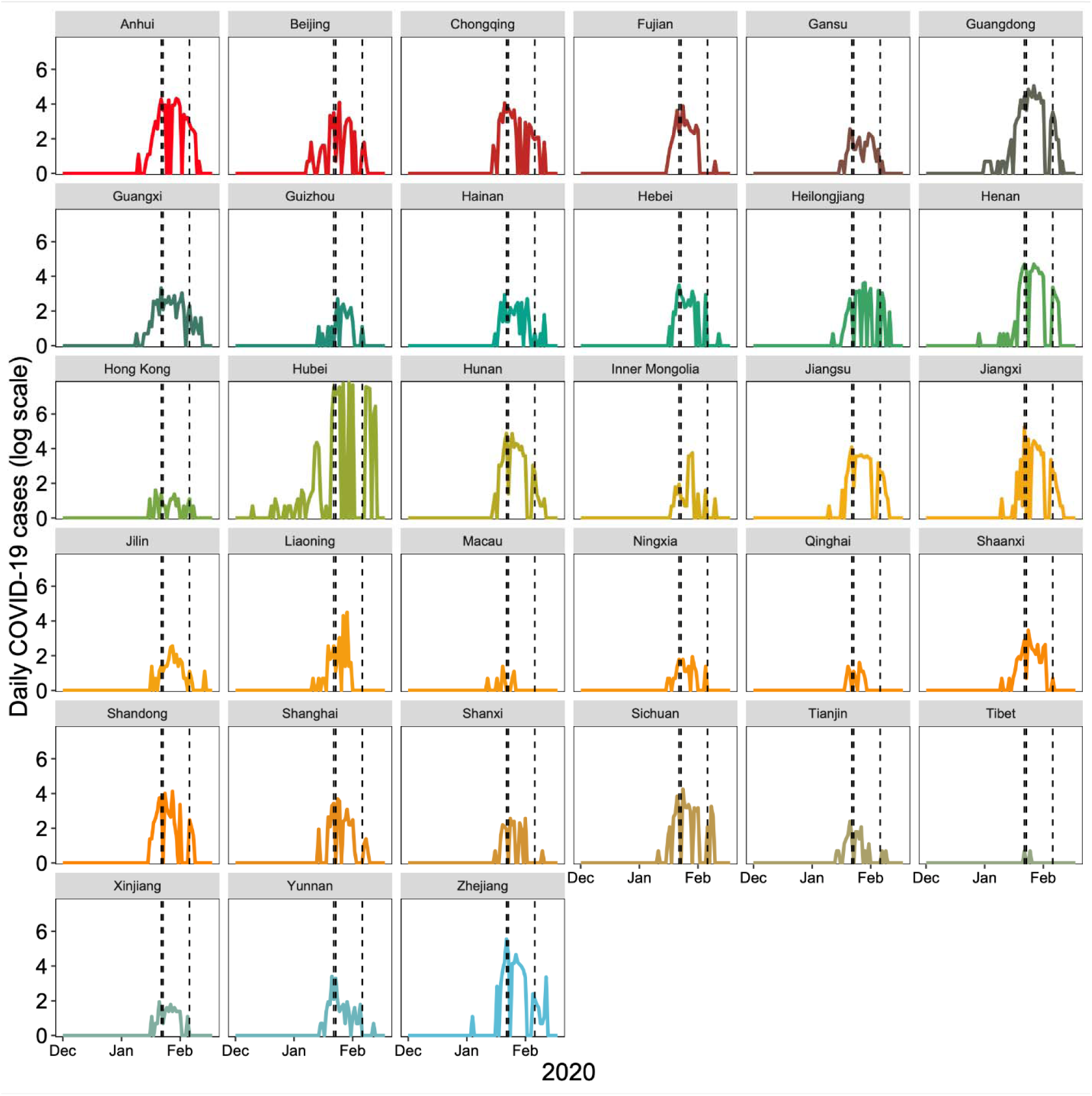
Daily case counts of COVID-19 in China between January 1^st^ and February 15^th^, 2020 (log scale).

**Figure S7:**
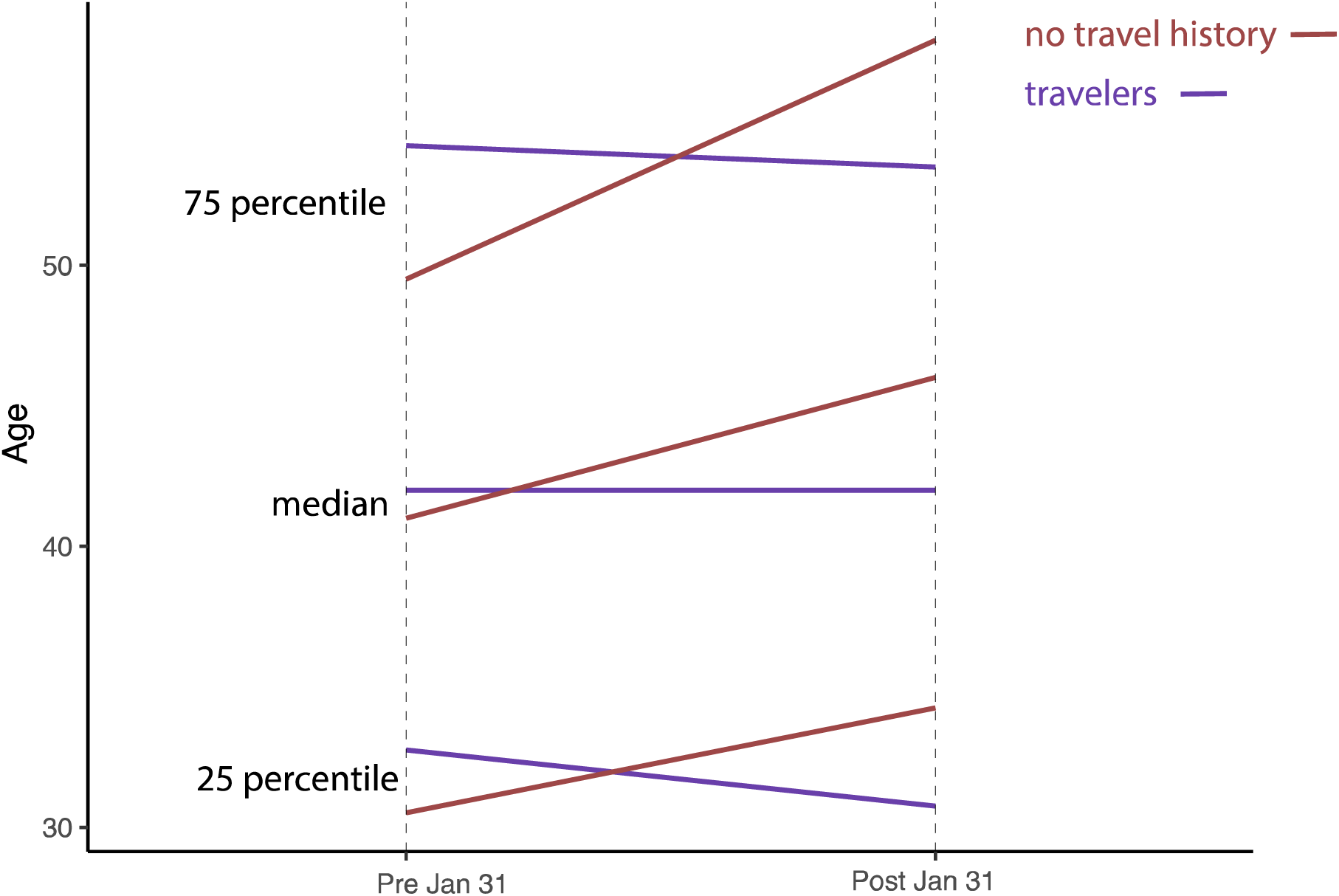
Median and 25, 75 percentiles of reported age of cases reporting travel history (purple) and those that do not (red) before 31 January 2020 and after.

**Figure S8:**
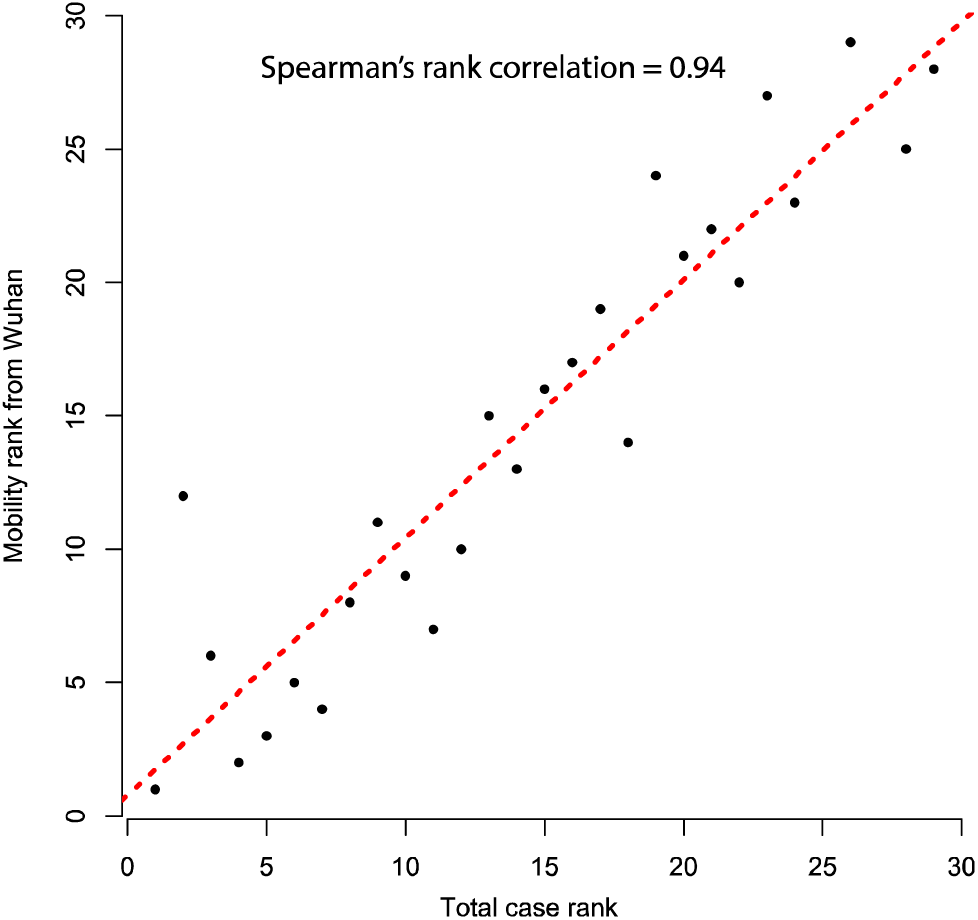
Correlation between total number of cases and human mobility from Wuhan.

## Members of the COVID-19 working group

Bo Xu, Sumiko Mekaru, Kara Sewalk, Lauren Goodwin, Alyssa Loskill, Emily Cohn, Yulin Hswen, Sarah C. Hill, Maria M. Cobo, Alexander Zarebski, Sabrina Li, Erin Hulland, Julia Morgan, Lin Wang, Katelynn O’Brien.

## References

1. S. Chen, J. Yang, W. Yang, C. Wang, T. Baerninghausen, COVID-19 control in China during mass population movements at New Year. Lancet (2020), doi:10.1016/S0140-6736(20)30421-9.

2. N. Zhu et al., A Novel Coronavirus from Patients with Pneumonia in China, 2019. N. Engl. J. Med., EJMoa2001017 (2020).

3. H. Tian et al., medRxiv, in press, doi:10.1101/2020.01.30.20019844.

4. Z. Du et al., Risk for Transportation of 2019 Novel Coronavirus Disease from Wuhan to Other Cities in China. Emerg. Infect. Dis. 26 (2020), doi:10.3201/eid2605.200146.

5. J. T. Wu, K. Leung, G. M. Leung, Nowcasting and forecasting the potential domestic and international spread of the 2019-nCoV outbreak originating in Wuhan, China: a modelling study. Lancet. 6736 (2020), doi:10.1016/S0140-6736(20)30260-9.

6. S. Cauchemez et al., Middle East respiratory syndrome coronavirus: Quantification of the extent of the epidemic, surveillance biases, and transmissibility. Lancet Infect. Dis. 14, 50–56 (2014).

7. J. Lessler et al., Incubation periods of acute respiratory viral infections: a systematic review. Lancet Infect. Dis. 9, 291–300 (2009).

8. J. A. Backer, D. Klinkenberg, J. Wallinga, Incubation period of 2019 novel coronavirus (2019- nCoV) infections among travellers from Wuhan, China, 20–28 January 2020. Eurosurveillance. 25, 20–28 (2020).

9. Q. Li et al., Early Transmission Dynamics in Wuhan, China, of Novel Coronavirus–Infected Pneumonia. N. Engl. J. Med., 1–9 (2020).

10. World Health Organization (WHO), Coronavirus disease 2019 (COVID-19) Situation Report – 27 (2020) (available at https://www.who.int/docs/default-source/coronaviruse/situation-reports/20200216-sitrep-27-covid-19.pdf?sfvrsn=78c0eb78_2).

11. Novel Coronavirus Pneumonia Emergency Response Epidemiology Team, The Epidemiological Characteristics of an Outbreak of 2019 Novel Coronavirus Diseases (COVID-19) — China, 2020. China CDC Wkly. 41, 145–151 (2020).

12. E. Goldstein, V. E. Pitzer, J. J. O’Hagan, M. Lipsitch, Temporally Varying Relative Risks for Infectious Diseases. Epidemiology. 28, 136–144 (2017).

13. World Health Organization (WHO), Novel Coronavirus (2019-nCoV) SITUATION REPORT - 3 (2020) (available at https://apps.who.int/iris/bitstream/handle/10665/330762/nCoVsitrep23Jan2020-eng.pdf).

14. J. F.-W. Chan et al., A familial cluster of pneumonia associated with the 2019 novel coronavirus indicating person-to-person transmission: a study of a family cluster. Lancet. 395, 514–523 (2020).

15. T. J. Hastie, D. Pregibon, in Statistical Models in S, J. M. Hastie, C. T. J., Eds. (Wadsworth & Brooks/Cole, 1992).

16. J. Riou, C. L. Althaus, Pattern of early human-to-human transmission of Wuhan 2019 novel coronavirus (2019-nCoV), December 2019 to January 2020. Eurosurveillance. 25, 1–5 (2020).

17. A. R. Tuite, D. N. Fisman, Reporting, Epidemic Growth, and Reproduction Numbers for the 2019 Novel Coronavirus (2019-nCoV) Epidemic. Ann. Intern. Med., 2019–2020 (2020).

18. Q. Li et al., Early Transmission Dynamics in Wuhan, China, of Novel Coronavirus–Infected Pneumonia. N. Engl. J. Med., EJMoa2001316 (2020).

19. M. J. Keeling, O. N. Bjørnstad, B. T. Grenfell, in Ecology, Genetics and Evolution of Metapopulations (Elsevier, 2004; https://linkinghub.elsevier.com/retrieve/pii/B9780123234483500192), pp. 415–445.

20. D. J. Watts, R. Muhamad, D. C. Medina, P. S. Dodds, Multiscale, resurgent epidemics in a hierarchical metapopulation model. Proc. Natl. Acad. Sci. 102, 11157–11162 (2005).

21. G. Chowell, L. Sattenspiel, S. Bansal, C. Viboud, Mathematical models to characterize early epidemic growth: A review. Phys. Life Rev. 18, 66–97 (2016).

22. C. Viboud et al., Synchrony, waves, and spatial hierarchies in the spread of influenza. Science. 312, 447–51 (2006).

23. M. J. Keeling, B. T. Grenfell, Individual-based Perspectives on R0. J. Theor. Biol. 203, 51–61 (2000).

24. K. P. Burnham, D. R. Anderson, Multimodel Inference. Sociol. Methods Res. 33, 261–304 (2004).

25. C. Fraser, S. Riley, R. M. Anderson, N. M. Ferguson, Factors that make an infectious disease outbreak controllable. Proc. Natl. Acad. Sci. 101, 6146–6151 (2004).

26. B. Xu, M. U. G. Kraemer, Open access epidemiological data from the COVID-19. Lancet Infect. Dis. 3099, 30119 (2020).

27. B. Xu et al., Sci. Data, in press.

28. R. E. Ramshaw et al., Adatabase of geopositioned Middle East Respiratory Syndrome Coronavirus occurrences. Sci. Data. 6, 318 (2019).

29. M. L. McHugh, The Chi-square test of independence. Biochem. Medica. 23, 143–149 (2013).

30. J. H. McDonald, Handbook of Biological Statistics (Sparky House Publishing, Baltimore, Maryland, ed. 3, 2014).

31. N. R. Faria et al., Genomic and epidemiological monitoring of yellow fever virus transmission potential. Science. 361, 894–899 (2018).

32. M. U. G. Kraemer et al., Past and future spread of the arbovirus vectors Aedes aegypti and Aedes albopictus. Nat. Microbiol. 4, 854–863 (2019).

33. G. Jin, J. Yu, L. Han, S. Duan, The impact of traffic isolation in Wuhan on the spread of 2019- nCov. medRxiv (2020), doi: https://doi.org/10.1101/2020.02.04.20020438.

34. A. S. Ai et al., Population movement, city closure and spatial transmission of the 2019-nCoV infection in China. medRxiv (2020), doi:doi.org/10.1101/2020.02.04.20020339.

35. D. Bates, M. Mächler, B. Bolker, S. Walker, Fitting Linear Mixed-Effects Models Using lme4. J. Stat. Softw. 67, 201–210 (2015).

36. J. Friedman, T. Hastie, R. Tibshirani, Regularization Paths for Generalized Linear Models via Coordinate Descent. J. Stat. Softw. 33 (2010), doi:10.18637/jss.v033.i01.

37. B. Ripley et al., MASS. R package version 7.3-51.5. (2019) (available at https://cran.r-project.org/package=MASS).

38. H. Heinzl, M. Mittlböck, Pseudo R-squared measures for Poisson regression models with over- or underdispersion. Comput. Stat. Data Anal. 44, 253–271 (2003).

39. A. Zeileis, C. Kleiber, S. Jackman, Regression models for count data in R. J. Stat. Softw. 27, 1–25 (2008).

